# APPLAUSE: Automatic Prediction of PLAcental health via U-net Segmentation and statistical Evaluation

**DOI:** 10.1101/2020.09.22.20199521

**Authors:** Maximilian Pietsch, Alison Ho, Alessia Bardanzellu, Aya Mutaz Ahmad Zeidan, Lucy C. Chappell, Joseph V. Hajnal, Mary Rutherford, Jana Hutter

## Abstract

**Purpose:** Artificial-intelligence population-based automated quantification of placental maturation and health from a rapid functional Magnetic Resonance scan. The placenta plays a crucial role for any successful human pregnancy. Deviations from the normal dynamic maturation throughout gestation are closely linked to major pregnancy complications. Antenatal assessment in-vivo using T2* relaxometry has shown great promise to inform management and possible interventions but clinical translation is hampered by time consuming manual segmentation and analysis techniques based on comparison against normative curves over gestation.

**Methods:** This study proposes a fully automatic pipeline to predict the biological age and health of the placenta based on a rapid (sub-30 second) T2* scan in two steps: Automatic segmentation using a U-Net and a Gaussian Process regression model to characterize placental maturation and health. These are trained and evaluated on 110 3T MRI placental data sets including 20 high-risk pregnancies diagnosed with pre-eclampsia and/or fetal growth restriction.

**Results:** Automatic segmentation achieves comparable performance to human experts (mean DICE coefficients automatic-manual 0.76, Pearson Correlation Coefficient 0.986 for mean T2* within the masks). The placental health prediction achieves an excellent ability to differentiate early cases of placental insufficiency before 32 weeks. High abnormality scores correlate with low birth weight, premature birth and histopathological findings. Retrospective application on a different cohort imaged at 1.5T illustrates the ability for direct clinical translation.

**Conclusion:** The presented automatic pipeline facilitates a fast, robust and reliable prediction of placental maturation. It yields human-interpretable and verifiable intermediate results and quantifies uncertainties on the cohort-level and for individual predictions. The proposed machine-learning pipeline runs in close to real-time and, deployed in clinical settings, has the potential to become a cornerstone of diagnosis and intervention of placental insufficiency.

## 1. Introduction

### A. Placental Maturation

The human placenta is key for any successful human pregnancy. It grows and changes across gestation to adapt to the increasing demands of the foetus. To this end the increased transfer of nutrients and oxygen from the maternal circulation to the fetal circulation is of key importance. This exchange process, occurring in 20-40 functional units or *lobules* across the placenta, relies on sufficient inflow from the maternal spiral arteries, through the fetal vasculature in the villous trees, into the umbilical vein, to match fetal demand. This low resistance system relies on ongoing combined angiogenesis and villous transformation with initial sprouting and branching followed by stromal reduction to increase the vasculo-syncitial membrane area to maximise oxygen and nutrient transfer. As the placenta ages, there is increased deposition of calcium within the lobules as well as deposition of fibrin largely within the septa between the lobules. The fibrin deposition contributes to the lobulated appearance on imaging, seen as low signal intensity on T2-weighted images, and the calcium deposition to an increase in granularity within the placenta as a whole. This process of placental ageing occurs normally in accord with gestational age (GA) and the associated changes are not pathological. However, accelerated aging of the placenta, as documented on histopathology, has been associated with increased risk of placental failure, fetal growth restriction, preeclampsia (PE) and unexplained late stillbirth (1, 2). In contrast, delayed maturation of the placenta is associated with gestational diabetes and chromosomal abnormalities (1–3). Thus, advancing our understanding of placental maturation and identifying tools to describe and quantify this process are vital to help improve early detection of placental failure. Functional MR imaging could provide a quantifiable marker of placental aging and could facilitate identification of accelerated ageing or of a delayed trajectory in individual placentas.

### B. State of the art

Placental development over gestation has been recently studied with T2* relaxometry (4–11). The T2* values can be linked to the concentration of deoxygenated haemoglobin via the BOLD effect and thus provide both an ability to visually inspect and quantify function in spatial maps cross-sectionally and over gestation. Many recent studies have employed these techniques to study the placenta in high-risk pregnancies compared to control low-risk pregnancies with normal outcomes. The majority of studies compare placental mean T2* values between controls and high-risk cohorts.

### Segmentation

A crucial step for any quantitative, or indeed, qualitative assessment is the detection and delineation of the placenta. The heterogeneous placental shape and variation of its location on images that typically cover the entire uterus, including maternal, fetal and placental tissue as well as amniotic fluid, hampers this step. For current published studies, the obtained T2* maps are manually segmented, either on selected slices (4, 6) or across the entire placental volume (7, 8, 12). First attempts to automate placental segmentation have already been undertaken for anatomical placental scans. The interactive Slic-Seg approach was proposed using random forests within slices (13) and subsequently improved using probability-based 4D Graph Cuts and deep learning. The latter utilizes user interactions after an initial convolutional neural network to refine the segmentation with a second convolutional neural network. Fully automatic frameworks were proposed using 3D multi-scale convolution neural networks to identify the area of interest, followed by 3D dense conditional random fields (14) and U-Net based segmentation (15). Finally, a technique combining motion correction, segmentation and shape extraction has been proposed by Miao et al. (16). Similarly, in 3D placental US imaging, seed-based random walker techniques (17) and U-Nets have been successfully employed for volumetric segmentations. However, for functional imaging data, to date only manual placental segmentations have been used (4, 6–8, 12, 18).

### Disease progression using Gaussian Processes

Conventionally, following segmentation, quantitative measures are obtained, most often the mean T2*, averaged across the entire placenta, which are assessed against “normative” curves over gestation obtained from longitudinal or cross-sectional studies (4, 6, 8, 19, 20). Using a group of placentas of normal appearance, a normative curve and credibility intervals can be derived directly from the data using a Bayesian regression algorithm. Assuming abnormal placental development manifests as a mean T2* signal deviation, this can be used to estimate an abnormality score, taking the uncertainty of the model fit into account. We use Gaussian Process regression (21), a nonparametric algorithm that assigns a probability to each possible function describing the training data, and allows the calculation of normative curves (the mean of its posterior distribution) as well as credibility regions around these. This technique has been successfully used previously, for instance, in the setting of neuroimaging (22–24). A key difference to these studies is however the inter-subject variability of placental shape, size and location which hinders the construction of atlases or standard planes. Hence, we use the scalar measures GA and mean T2* inside the automatically segmented placenta to estimate normal and abnormal maturation.

A placental age prediction model could be used to identify abnormal data via comparison of ‘predicted biological placental age’ and chronological age. Similarly, we could predict mean T2* for a given GA and determine the difference between measured and expected values. We use an approach based on error-in-variables modelling, that takes uncertainties associated with both quantities into account to minimise prediction bias.

### C. Contributions

This work seeks to establish a fully automatic pipeline using mean T2* measurements as a biomarker of placental maturation and health. A data set of well characterized uncomplicated pregnancies as well as a range of complicated pregnancies allows us to study placental maturation in depth. We aim to do this in two different ways:

1. Demonstrate the automated pipeline consisting of a sub-30 second MRI scan, an automated U-Net based segmentation, and subsequent placental health estimation.
2. Demonstrate that the studied techniques can accurately capture the credibility interval of low-risk data which can be used to build a normative model of placental maturation in a cohort of well characterized uncomplicated pregnancies and to assess a cohort of pregnancies affected by pregnancy complications.

## 2. Materials and Methods

The proposed pipeline is depicted in Fig. 1 consisting of the data acquisition, automatic segmentation used to calculate the placental mean T2* and the Gaussian Process regression fit or prediction, which is used to characterize placental health. All parts will be discussed in detail in the following.

**Fig. 1.**
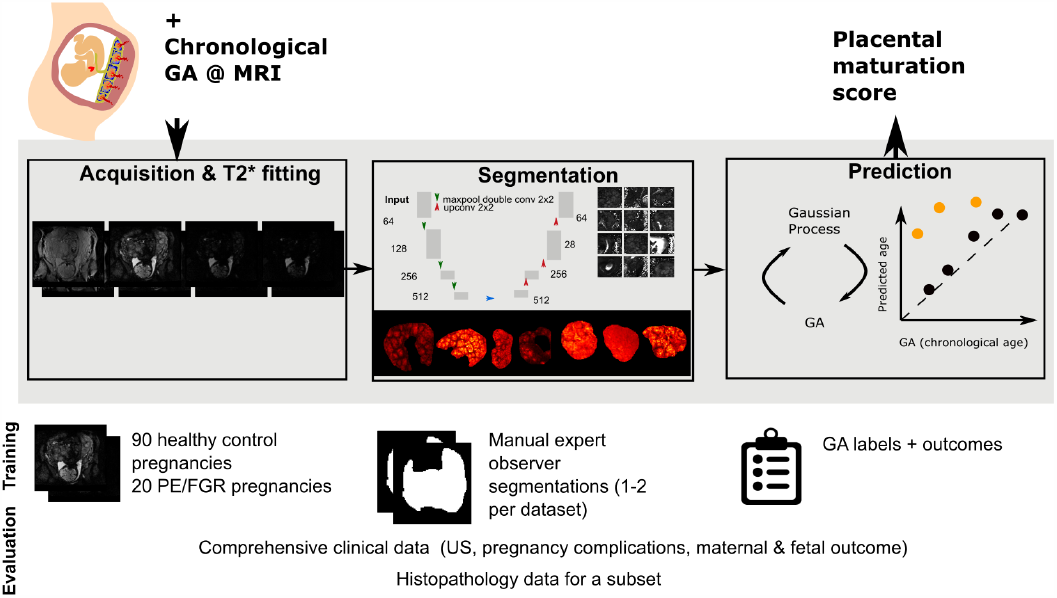
The proposed algorithm is presented in the grey box together with input and output above, training data and evaluation data below.

### A. Data acquisition and preparation

MRI imaging was performed on 108 women with singleton pregnancies between 18 and 40 weeks GA without contraindications to MRI. Informed consent was obtained (https://placentaimagingproject.org/, REC 14/LO/1169) and the scan was performed on a clinical 3T Philips Achieva (Best, The Netherlands) scanner using the 32-channel cardiac receiver coil. All women were scanned in the supine position with frequent monitoring of their heart rate, saturation and blood pressure throughout the scan. Dedicated padding was provided to increase maternal comfort and verbal communication was maintained. Clinical information and pregnancy outcome were obtained. For the fetus, this included gestation at birth, birth weight and birth weight centile calculated using the INTERGROWTH algorithm (25), Apgar score and admission to neonatal unit. Maternal age, Body Mass Index (BMI), clinical history, ethnicity and any diagnosis of PE, fetal growth restriction, hypertension, gestational diabetes mellitus and any other complications were recorded. Analysis of subsequent clinical outcome data allowed us to define a low-risk cohort and a high-risk cohort diagnosed with PE or growth restriction using the criteria outlined previously (26, 27). The GA at scan was obtained based on the agreed expected date of delivery from ultrasound exams between 11 and 14 weeks of gestation. In addition, for 39 participants of the healthy and 17 participants from the high-risk cohort histopathological data was available. Macroscopic and microscopic evaluation of the placenta was performed and the presence of maternal vascular malperfusion and signs of chorioamnionitis were recorded. Details of these cohorts are given in Table 2 and graphically illustrated in Supplementary Figure 8.

**Table 1.** MRI parameters.

|  |  |
| --- | --- |
| **FOV** [mm × mm] | 340×400 |
| **Resolution** [mm<sup>3</sup>] | 3×3×3 |
| **Slices** | 24-86 |
| **Orientation** | coronal |
| **TEs** [ms] | 13.8, 70.4, 127, 183.6 |
| **TR** [s] | 12 |
| **SENSE** | 3 |
| **Halfscan** | 0.6 |
| **TA** [s] | 26 |

**Table 2.** Characteristics of the study participants. The cohort included low-risk pregnancies without evidence of PE, fetal growth restriction, Gestational Diabetes Mellitus or hypertension resulting in a live birth at 36 weeks or above with a birth weight between the 2nd and 98th centile and a high-risk cohort of pregnancies diagnosed with PE and or fetal growth restriction. Δ GA corresponds to the time between scan and delivery in weeks. For scatter plots see Figure 8.

|  | Low-risk | Placental insufficiency |
| --- | --- | --- |
| **n** | 90 | 20 |
| **GA at MRI** | 29.93 ± 4.26 | 30.83±3.52 |
| weeks |  |  |
| **Maternal age** | 33.9 ± 3.54 | 32.52 ± 6.69 |
| years |  |  |
| **BMI** | 22.48 ± 2.66 | 24.65±2.99 |
| kg/m<sup>2</sup> |  |  |
| **GA delivery** | 40.09±1.23 | 34.07±3.27 |
| weeks |  |  |
| **Fetal sex** | 40.09 | 34.07 |
| % female |  |  |
| **Birth weight** | 3.41 ± 0.43 | 1.77±0.7 |
| kg |  |  |
| **Birth weight** | 55.72±27.28 | 13.45±18.39 |
| centile |  |  |
| **Δ GA** | 10.06±4.46 | 3.23±3.27 |
| weeks |  |  |

After initial fetal brain imaging and whole uterus imaging, a map of the B0 field was acquired and image-based shimming performed. Next, a free-breathing multi-echo Gradient Echo Echo Planar Imaging technique was performed, the parameters are specified in Table 1. The local requirement is to keep the acoustic output of all sequences used for routine fetal imaging below 98 dB(A) which drives the choice of TEs and resolution. The total acquisition time was 26 seconds. Of the 108 acquired scans, 6 data sets with either signs of sub-clinical contractions, seen as transient areas of low signal distorting the uterine wall, or containing unresolved fat artefacts were discarded. This data pre-selection step can be performed in less than one minute by trained placental analysts. Work on automatic identification based on 3D reconstruction is ongoing (28).

A mono-exponential decay model was fitted on a voxel-level using a Levenberg-Marquart algorithm with 50 iterations, initialised using the first echo time. Slices were fitted independently, and as all echos for one slice were acquired within 200ms, no motion was observed *within slices* obviating the need for motion correction. T2* values over 300ms were clipped to limit partial volume effects from amniotic fluid close to the placental boundaries.

The placenta was manually outlined on all slices by one or two experienced placental analysts using the fitted T2* map and/or the image data corresponding to individual echo times. If two segmentations were available, one was randomly chosen and used for training and evaluation of the U-net. The strict selection of normal, uncomplicated pregnancies forming the normal cohort allows using the chronological GA as a substitute for biological placental age.

### B. Placental segmentation

The network architecture is based on the 2D U-net implementation *nnU-Net3* (29). Training and inference were performed on 2D patches of 64×64 voxels. The data pre-processing included normalizing of the T2* maps by demeaning with the mean of the training dataset. For training, data augmentation in the form of spatial cropping, mirroring and rotations was applied using the *batchgenerator* framework^1^ (30). The network is trained via stochastic gradient descent using the Adam optimizer (31) with a learning rate of 0.0001 and batch size of five for 50000 epochs using a binary cross entropy loss function. The training of the network utilizes 12 GB of VRAM and takes approximately 2 days on a Tesla P100 GPU on Google collaboratory.

### Training and test data for the segmentation

To test out-of-sample performance, five-fold stratified cross-validation was performed. All reported segmentation and hereof based T2* measurements were generated using the network for which this data was part of the held-out data. To avoid leakage between training and test data, participants were assigned exclusively to one of the partitions. Each training round was performed on 72 healthy low-risk cases and 16 high-risk abnormal cases, the test data consisting of 18 control and 4 high-risk cases. If two segmentations were available, one was chosen randomly to perform the training as specified above.

### Evaluation

Each network was evaluated on its corresponding test data. To match the training process, inference was performed on 64×64 patches of the validation set with patches chosen to overlap by 30%. The predicted likelihood was averaged across overlapping predictions and binarized to obtain the full 3D mask. The performance was evaluated with two metrics, first via direct evaluation of the segmentation using the DICE coefficient (32). Second, agreement between the mean T2* value between the manual and automatic segmented placental mean T2* results was evaluated on the test data using the Pearson Correlation Coefficient and Root Mean Square Error.

### C. Unbiased Placental health prediction

The final part of the pipeline consisted of a Gaussian Process regression model that uses the mean T2* data generated via automated segmentations to capture normal maturation.

### Biological age prediction

To predict biological placental age *t*_*b*_ from the mean T2* value inside the automatically segmented placental mask 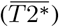, we assumed a probabilistic function 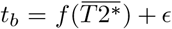 with independent and identically normally-distributed noise 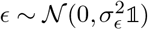. We trained a Gaussian Process regression model to approximate 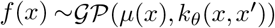, where *µ*(*x*) is the mean function and *k* the covariance function with hyperparameters *θ*. Given the relatively smooth and continuous relation of the data in a GA-vs-T2* plot, we chose *µ* to be constant and *k* to model a linear trend with local nonlinear deviation using a sum of dot product kernel and squared-exponential function kernel (length-scale ≤ 1*σ*(*T* 2*\**)) with an additive white noise kernel. Model hyperparameters were estimated by maximizing the log marginal likelihood, maximizing the probability of the training data conditioned on kernel hyperparameters. All fits were performed using *SciKitLearn* (33). The performance was evaluated by calculating the mean squared error and Pearson correlation coefficient between biological GA (*t*_*b*_) and predicted GA.

### Total least squares Gaussian Process regression

In the ordinary least squares regression model 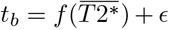, it is assumed that the independent variable 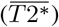 is measured without error; all error is attributed to *t*_*b*_. However, 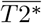 values are derived from noisy MRI measurements using imperfect segmentations and 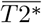 might be influenced by factors not related to age. Since biological age is known relatively precisely, the majority of the errors in the least squares fit could be associated with the independent variable. To obtain an unbiased estimates of placental health, an errors-in-variables model (34–37) that takes both sources of error into account is required. A commonly used errors-in-variables technique is total least squares, which minimises the squared distance orthogonal to the fitted line or to the fitted (hyper-)plane in higher dimensions instead of along the direction of a single dependent variable. Different contributions to the expected residuals can be accounted for by scaling the data prior to fitting so that their expected errors are equal (36).

To be able to use a Gaussian Process regression fit, we transformed each of the 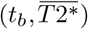 data points to a form that allowed fitting an approximately unbiased estimator using a least squares fit. For this, the age and mean T2* of the *n* measurements are stored in the columns of the 2 × *n* data matrix *X*, which is demeaned and, as in the TLS approach, scaled according to the expected errors in the training set of both quantities *X*_*s*_= *SX*, with 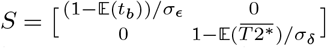. Finally, *X*_*s*_ is rotated so that its rows represent the scaled data transformed onto the first and second principal axes *X*′ = *U* ^*T*^ *SX*, where the columns of *U* hold the left singular vectors of *X*_*s*_.

### Training and test data for the prediction

We aim to evaluate the performance of the proposed pipeline on as much data as possible. Since the Gaussian Process regression model relies on the automatic segmentation, we used the segmentations generated on the cross-validation test-set and the respective U-net models to predict placental masks. This allowed calculating unbiased mean T2* values for all images. For training and evaluation of the Gaussian Process based health prediction, data from the low-risk control cohort were split into a training and test set in the ratio 0.7:0.3. Only low-risk cases were chosen for training as only these allow the Gaussian Process to accurately model normal maturation over gestation. The model was tested on the hold-out low-risk test data and the data from the high-risk cohort.

### Evaluation

A Gaussian Process regression fit was performed to predict the first row of *X*′, given the second. This allows capture of any potentially non-linear relations not accounted for by the first principal direction and to determine Z-scores and confidence regions using the Gaussian Process posterior. Z-scores for unseen data can be estimated using the transformation matrices and Gaussian Process posterior estimated from the training data. For visualisation, Gaussian Process predictions can be transformed back and shown in the original space of *X* using the inverse projection *S*^−1^*U*.

### Likelihood for accelerated ageing

The posterior predictive distribution of a Gaussian Process regression model can be used to describe the predictive distribution which allows estimating credibility intervals of the data. This allows the calculation of Z-scores for new observations. The posterior is conditional on the training data and on the hyperparameters but it assumes noiseless input data for inference. Given knowledge of the uncertainty associated with the input data, we can integrate over this input data distribution to estimate the true predictive distribution for noisy data. We assume a multivariate normal noise model with covariance matrix diag 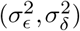 for new observations and estimate the corresponding predictive distribution via Markov Chain Monte-Carlo sampling (10000 samples). Conditional on the hyperparameters, training data, and input noise estimates, we can then calculate the likelihood that a placenta exhibits abnormally accelerated aging (*Z* < −3) from a single mean T2* measurement.

### D. Generalization and 1.5T data

Summarizing, once trained, the complete algorithm for unseen data consists of the acquisition and mono-exponential fitting of multi-echo gradient echo data followed by segmentation via the trained U-net and finally estimation of the Z-score or accelerated aging probability using the Gaussian Process Regression model. The translation to a new scanner and MRI protocol requires retraining or fine-tuning the segmentation U-net and estimation of the Gaussian Process parameters. Hence, translation depends upon a cohort of low-risk scans spanning the age-range of interest as well as manual segmentations for low-risk and, depending on image characteristics, high-risk groups.

To investigate generalization of the methods to other clinical cohorts and scanner environments, data was acquired on a clinical 1.5T Philips Ingenia scanner on cohorts of women with low-risk and high-risk pregnancies. These cohorts were in general characterized by a higher BMI due to the larger bore size of the employed 1.5T scanner. We note that increased maternal weight is associated with decreased image quality and increased maternal health risks including PE (38). The high-risk 1.5T cohort was uniquely composed of PE cases. A total of 42 data sets were acquired, including 36 control and 6 PE cases. The protocol was adapted as far as required for the different scanner conditions. Specifically, the inter-echo spacing was changed to adapt to the acoustic output of the 1.5T scanner. The spatial resolution was slightly increased from 3mm to 2.5mm to comply with other ongoing projects. This data was processed and analysed using the pipeline developed for the 3T data as described in the method section.

## 3. Results

We present results for all steps of the pipeline in the order of the processing steps. First, representative T2* data sets obtained with the sub-30 second whole placenta acquisition are illustrated in Fig. 2 ordered by GA. Second, the results of the segmentation step are presented and quantified and finally, the placental health prediction step is evaluated.

**Fig. 2.**
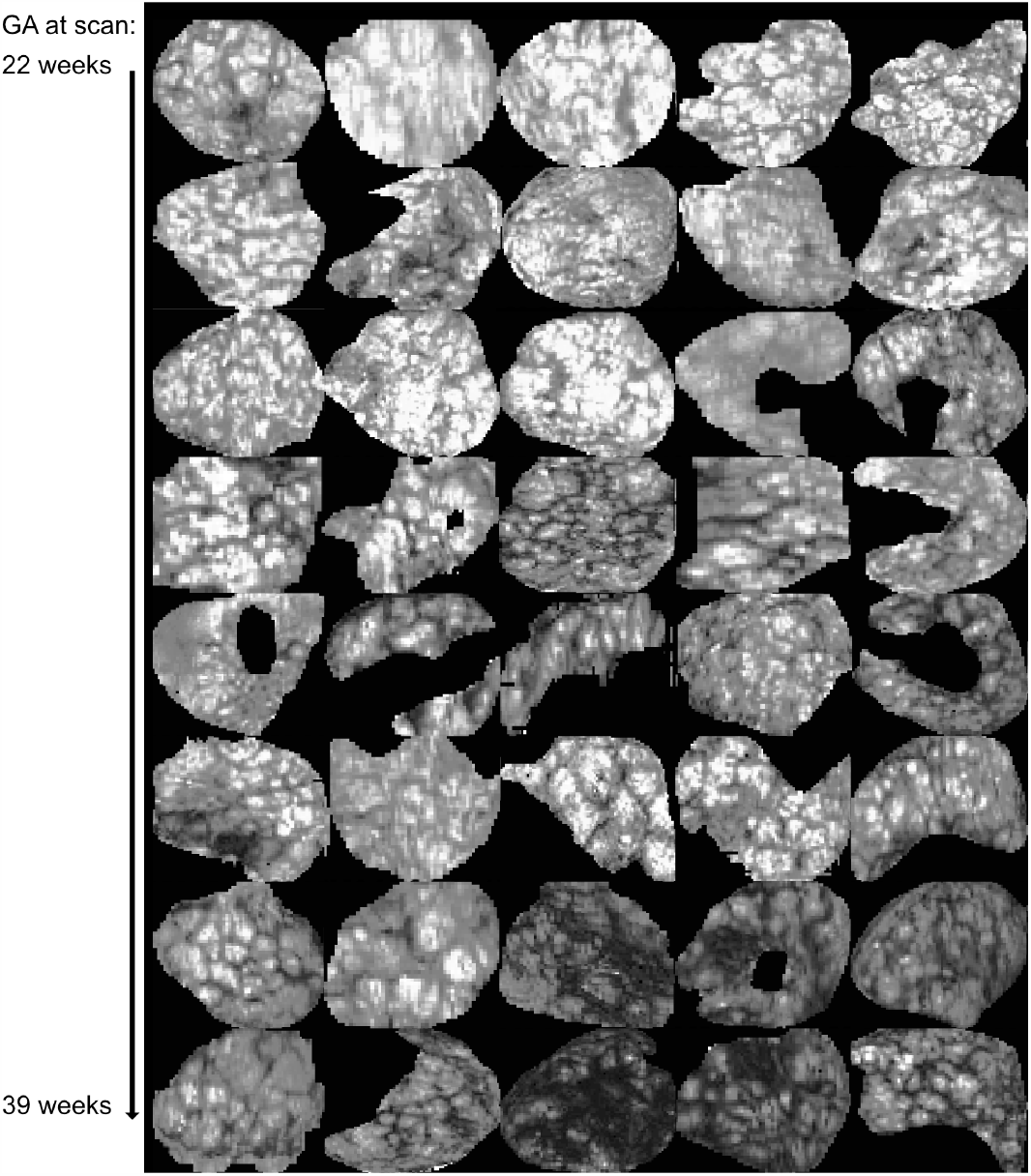
Illustration of central slices from segmented placental T2* maps for 40 low-risk cases used in this study. Images are sorted by GA (row-major).

### A. Segmentation

To evaluate the performance of the automatic segmentations, we compared the U-net segmentations obtained on the test sets of the cross-validation training scheme against one (randomly chosen) human expert segmentation. Automatic segmentations for two exemplary placentas are shown in Fig. 3, one with a median DICE score between manual and automated segmentations and one with a DICE score in the lowest 20%. The manual segmentations are shown for both observers in blue and green and the result of the automatic segmentation in red for two cases. Fig. 4 shows a quantitative comparison of segmentation results (DICE scores) and agreement in derived mean T2* values for the automatic segmentation compared to a randomly chosen manual segmentation and for the two expert annotators where data from both was available. On average, the agreement between human annotator measured as the DICE coefficient is comparable to the agreement between automatic and manual segmentation (c-d). The mean DICE coefficient for manual and automatic segmentation methods is 0.72 and 0.78, respectively. The variance between human and automated segmentations tends to be higher than that across the human observers. However, there is a good correspondence of the mean T2* results between automatic and manual segmentations (a) and between expert annotators (b) indicating that segmentation performance is sufficient to derive meaningful T2* values. The Pearson correlation coefficients for inter-rater and automatic segmentation performance is 0.987 and 0.884 respectively and the root mean squared error equals 3.2 and 7.4 ms respectively.

**Fig. 3.**
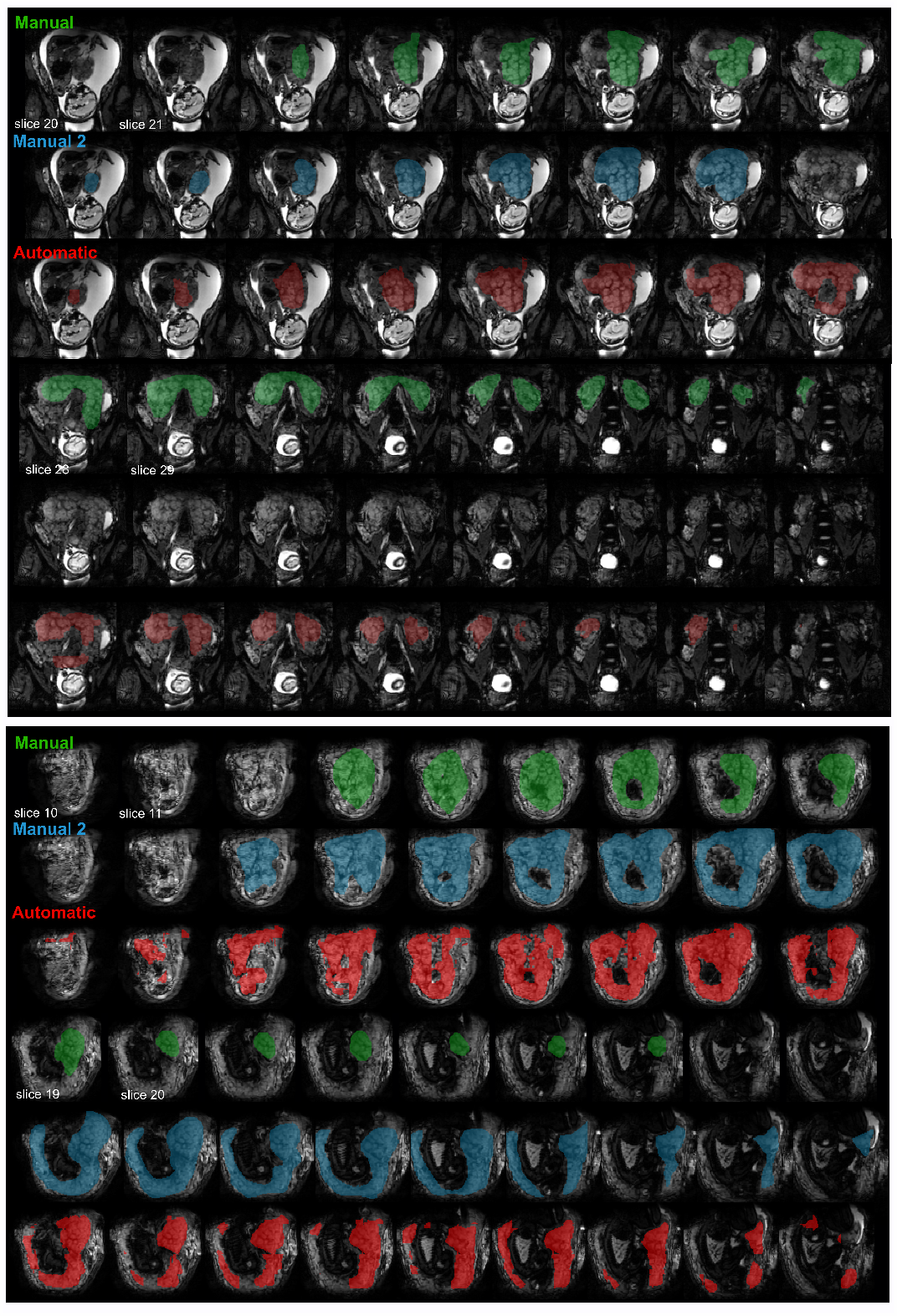
Segmentations from both manual (green) and automatic (red) process for two cases with Dice scores between automatic and manual segmentation in the lowest 20% (top) and highest 20% (bottom) of the normal cohort are illustrated overlaid on the volume acquired at the second TE. The slices are acquired in coronal plane from anterior to posterior position. In the top example, the ‘Manual 2’ segmentation includes only the first seven slices of the placenta, which explains the low Dice score here.

**Fig. 4.**
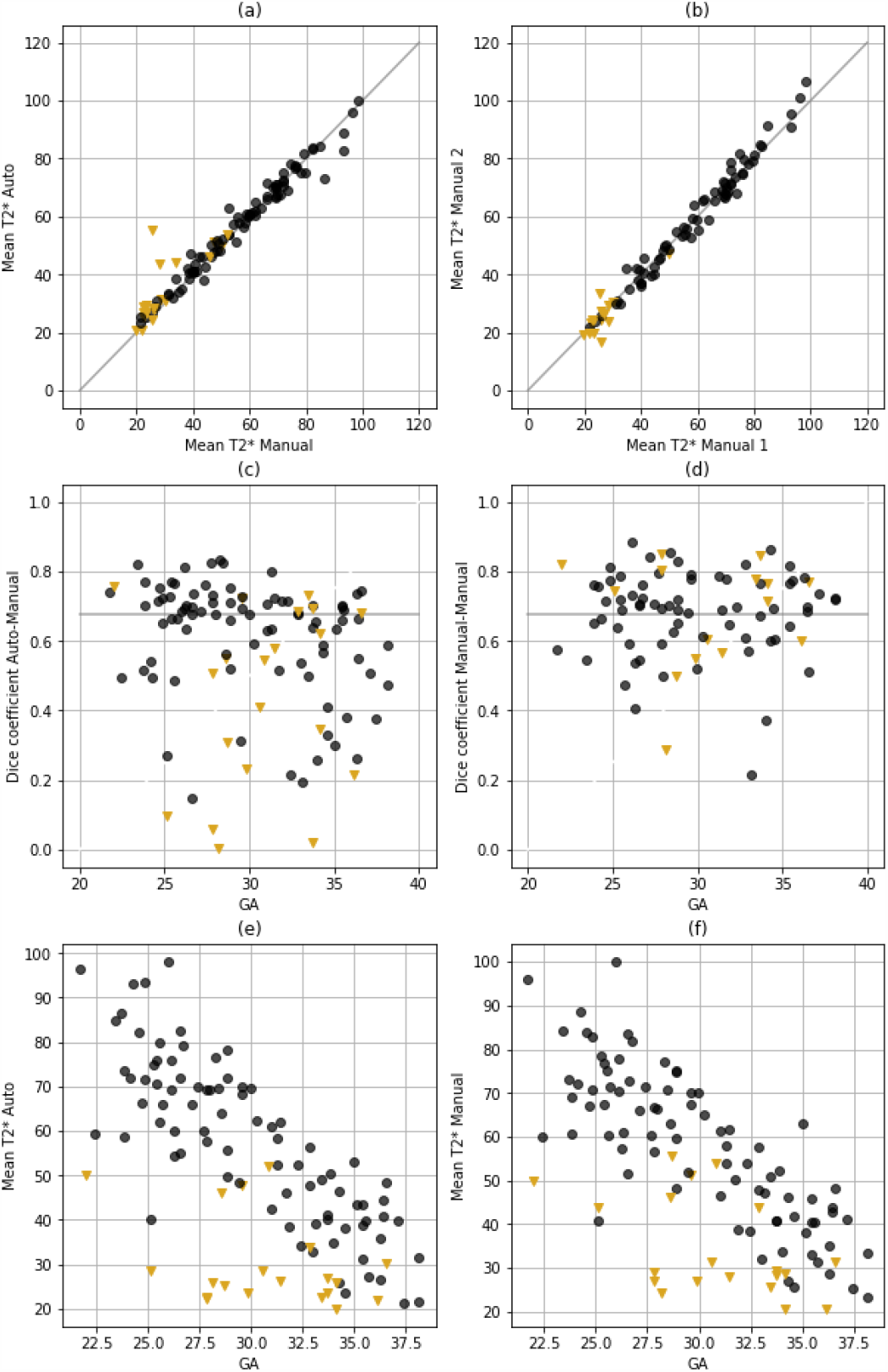
Quantitative results for the automatic segmentation. The results from the automatic segmentation in comparison with the one randomly chosen manual segmentation are given in the left column (a,c,e) and the results from the manual observers in the right column (b,d,f). (a) and (b) show the correspondence in mean T2* with the gray identity line. Plots (c) and (d) compare the Dice coefficient for all participants and (e) and (f) the mean T2* over GA. The gray line in (c-d) illustrates the mean Dice coefficient obtained between manual segmentations. As before, the results from the normal cohort are shown in black and the results from the abnormal cohort with mustard triangles. Units: GA in weeks, T2* in ms.

We further visually assessed the manual segmentations in detail. Differences, where present, were usually detected at the placental-myometrium boundary or in scans with reduced image quality due to maternal habitus or posterior located placenta.

### B. Placental health prediction

#### Data uncertainty estimate

For the total least squares fit, we estimate the errors associated with the two variables of interest. We model biological placental age using GA, which is not necessarily an absolute measure of development as normal full term gestation ranges from 37 to 42 weeks. Therefore, assuming Gaussian distributed errors, we expect the standard deviation of the biological age of the placenta *σ*_*E*_ to be (42−37)*/*(2 1.96) 1.3 weeks. We assume that the true not observable value of 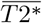 is corrupted by additive errors 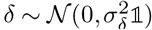 With fixed *σ*_*E*_, we estimate the 95% CI for 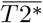 from a point estimate in 1*σ* age proximity of the highest data density as 35*ms*, yielding *σ*_*δ*_ = 8.7*ms*, which is slightly larger than 7.4*ms*, the root mean squared error of mean T2* between the manual and automatic segmentations. The 1.5T data consists of fewer measurements which hinders robust uncertainty estimation. Therefore, we pooled all low-risk data (training and test) to define four age bins containing at least 4 measurements within 1±*σ*_*E*_ weeks which was assumed to be the same for this cohort. *σ*_*δ*,1.5*T*_ = 16.4*ms* was estimated by averaging the T2* standard deviations of these age bins.

#### Placental biological age and health prediction

The total least squares Gaussian Process regression model (transformed back into GA vs mean T2* space) shows the expected approximately linear relationship between the two quantities (Fig. 5 (a)). Both training and test data lie within the expected Z-score range ± 3 (Fig. 5 (b)), indicating that data from the normal cohort is indeed modelled as such. Furthe more there is no discernible age-dependency in the Z-scores of the normal cohort. This is a result of the total least squares fit as demonstrated in Supplementary Fig. 9, where the Z-scores of an ordinary least squares model that was fit to predict biological placental age from mean T2* measurements shows a clear age-dependency in the training and test data (Supplementary Fig. 9 (b)). This age-bias of the ordinary least squares model causes erroneously elevated Z-scores for high-GA cases which reduces predictive power of accelerated aging in the high-risk cohort (see bottom of Fig. 6).

**Fig. 5.**
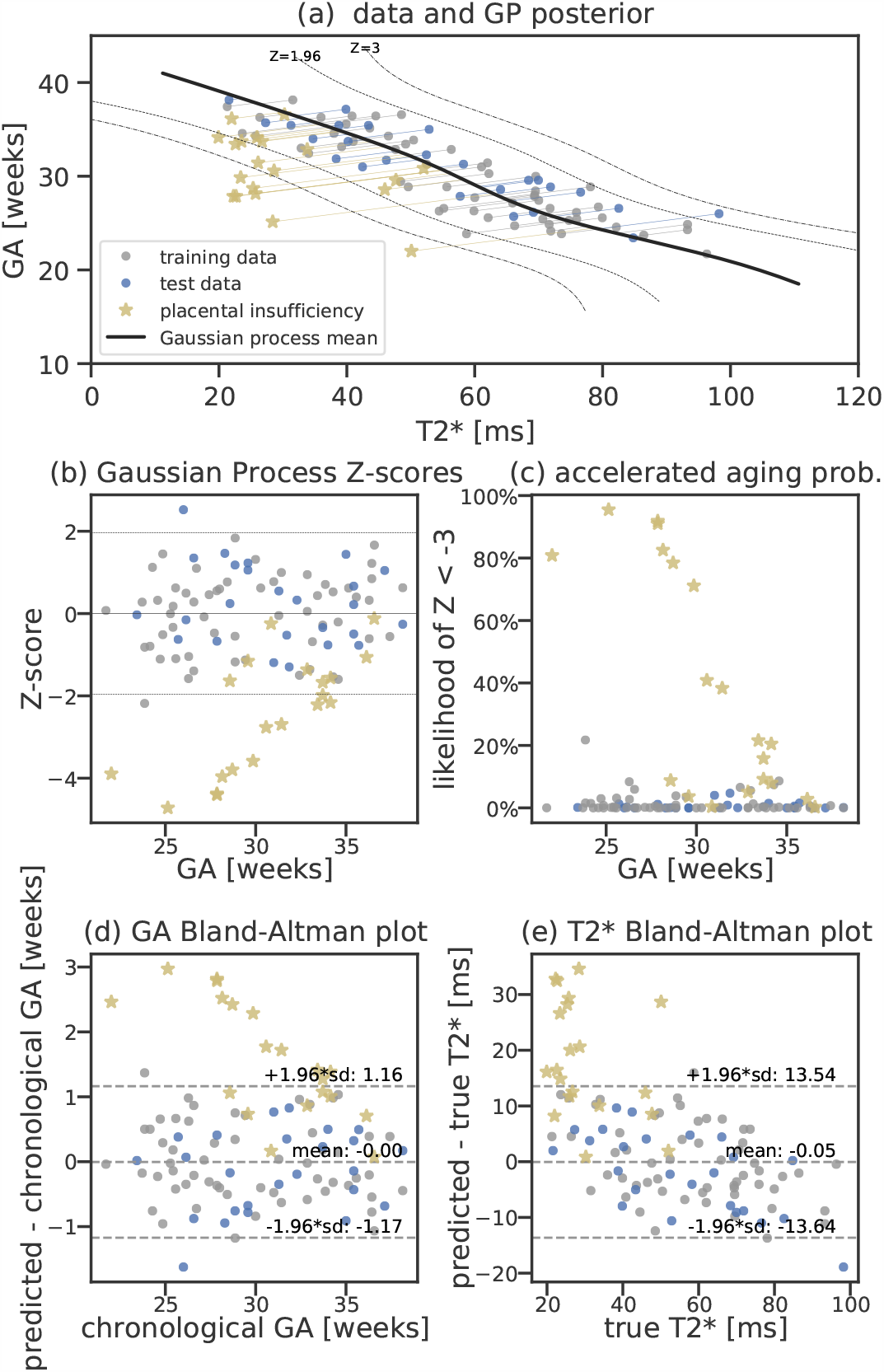
Total least squares Gaussian Process placental health prediction results. (a) Posterior mean and 95% (Z=±1.96) and 99.7% (Z=±3) credible intervals (dashed lines) estimated from the training data (gray). Test low-risk data and the high-risk data are shown in blue and yellow. Residuals are indicated using lines projecting data onto the posterior mean. (b) Scatter plot of Z-scores vs gestational age show no age bias in training and test data, high-risk cases tend to have reduced Z-scores indicating lower mean T2* or lower biological GA than expected. (c) Likelihood of accelerated aging for each observed data point taking the Gaussian Process credibility interval as well as uncertainty estimates of biological GA and mean T2* measurements into account. (d-e) Bland-Altman plots of age and mean T2*. Note that these plots are not independent as the prediction links both quantities. For least-squares fit and Gaussian Process results based on manual segmentations see Supplementary Figures 9 and 10.

**Fig. 6.**
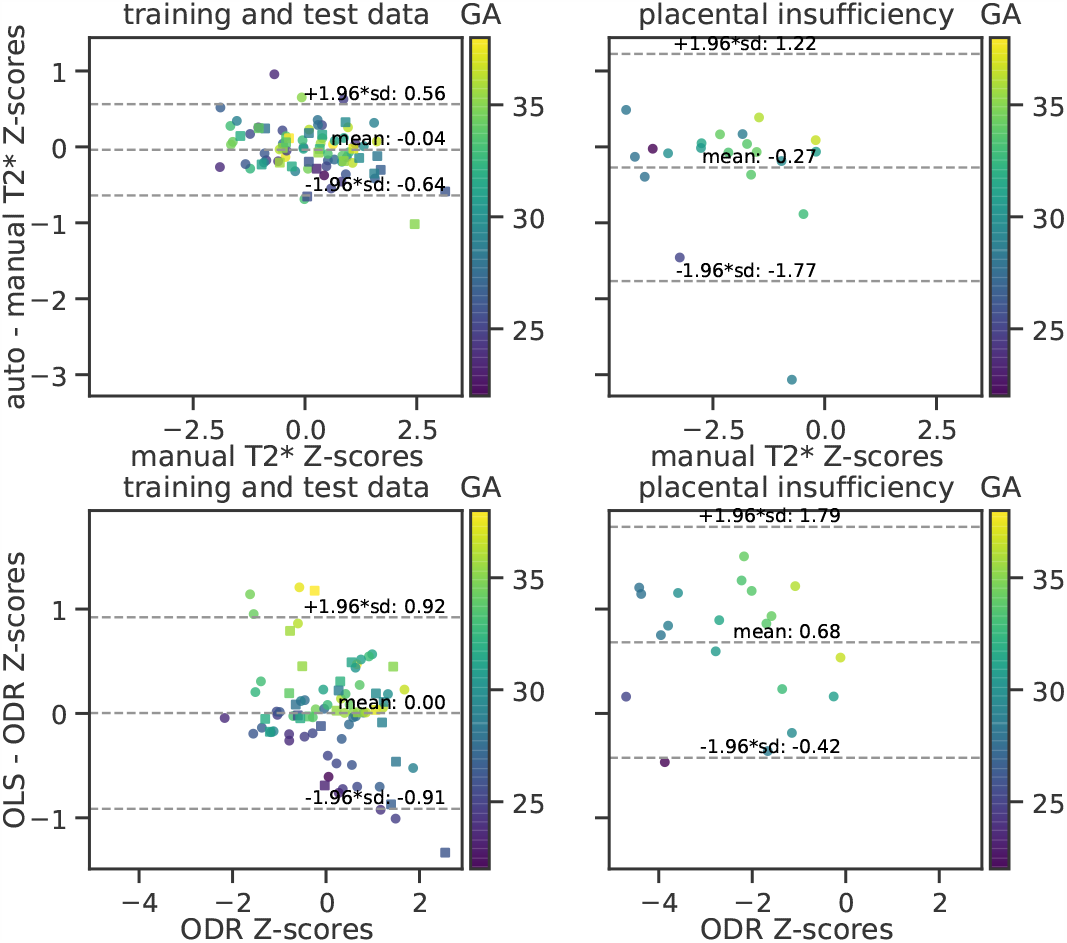
Top: Comparison of the obtained Z-scores between manual and auto-matic segmentation results both using total least squares regression illustrated with Bland-Altman plots for the low-risk cohort (left, training data shown as squares) and the high-risk cohort (right). Bottom: Bland-Altman plots for total least squares and ordinary least squares Gaussian Process fits using automatically generated masks. The GA at scan (in weeks) is color coded.

Utilizing a total least squares model, trained and evaluated on either manual or automatic segmentations, yields comparable predicted Z-scores for the low-risk cohort (±1.96*SD*[−0.64, 0.56]) and no discernible age-bias (see Bland Altman plots in Fig. 6). The abnormal cases exhibit similar bias and spread except for two to three cases, for which automatic segmentation-based Z-scores indicate higher abnormality than obtained via the manual segmentation. Hence, the predictions using automatic segmentations yield similar performance in the low-risk cohort and increase the rate of detected accelerated aging in the high-risk cohort.

#### Z-score correlates with biological and histopathological information

The obtained Z-scores are further analyzed and depicted in Fig. 7. A positive linear trend between Z-scores and both GA at birth and the logarithm of the birth weight centile can be observed in (a). Large deviations from the trends (Z-score < 4) are associated with either premature birth (GA at birth 28 weeks) or very low birth weight centile (< 1st centile). Correlation between Z-score and available histopathology findings are shown in Fig. 7 (b). In high-risk cases, lower Z-scores are associated with maternal vascular malperfusion (MVM) while signs of chorioamnionitis coincides with normal Z-scores. No cases with MVM have been found in the analysed low-risk cohort, and the Z-scores of the cases with chorioamnionitis findings in the low-risk cohort obtain similar (normal) Z-scores.

**Fig. 7.**
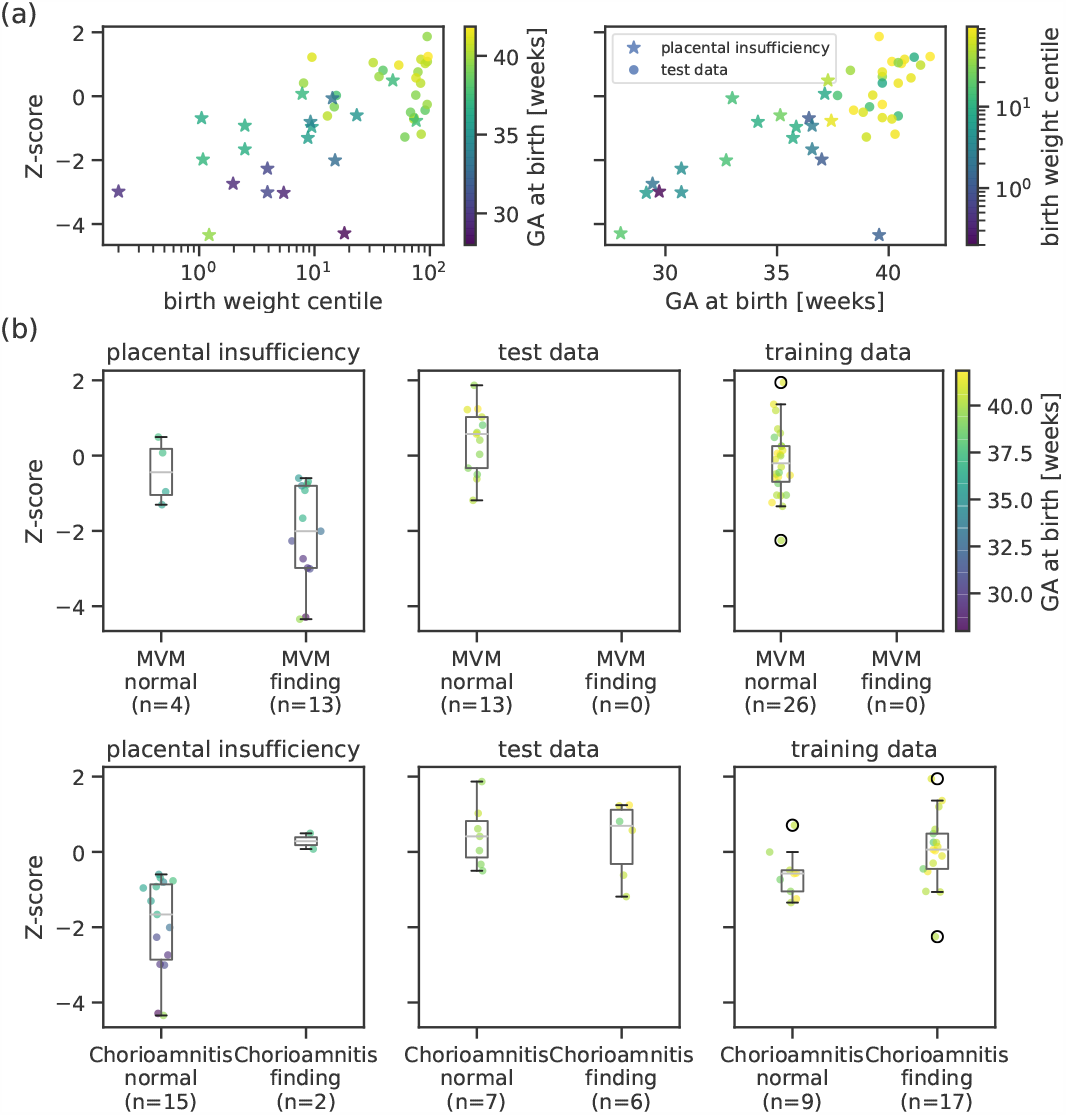
Top: The placental health Z-scores for the high-risk cohort and the low-risk test data are strongly related with degree of prematurity measured as GA at birth (right) and extremely low birth weight centile (left). Bottom: Z-scores grouped by histopathological examination results for training, test, and placental insufficiency data, color-coded by GA at birth. Participants with maternal vascular malperfusion (MVM) exhibit lower Z-scores and belong to the group delivered prematurely. Chorioamnionitis does not affect Z-scores in control cases but seems to correlate to normal Z-scores in two of the high-risk participants.

#### C. Generalization and 1.5T data

The results from the 1.5T data in Supplementary Figure 11 illustrate the same relationships between placental parameters and clinical findings as for the 3T data. The subjects with PE have consistently reduced Z-scores (*Z* < −1.5, Fig. 11(b)) and elevated likelihood of abnormally accelerated aging (Fig. 11(c)).

## 4. Discussion and Conclusion

This study presents a fully automatized pipeline to assess the maturation of a placenta in-vivo from a sub-30 second MRI scan. The proposed two-step process, consisting of first a fully automatic segmentation and second the assessment of placental age and health, assures accessibility and interpretability. The availability of the whole organ segmentations as an intermediate step allows visual inspection and effortless extension to other quantitative markers. Its potential for direct translation is illustrated by including results from a different cohort, scanned on a different scanner with lower field strength and altered acquisition parameters (Fig. 11.

The mean T2* values obtained from the automatic segmentations (Fig. 4) decay over GA in line with literature values (4, 9, 12, 18), and are reduced as previously observed in high-risk cases (19, 26). The obtained placental Z-scores for the high-risk cohort with apparent placental insufficiency correlate well with outcome assessed by birth weight centile, GA at birth and histopathological results (Fig. 7). In addition, reduced Z-scores, hypothesized to relate to accelerated aging, corresponded to the presence of maternal vascular malperfusion in this study. This is in line with the most common components of MVM, villous infarction, retroplacental hemorrhage, accelerated villous maturation, and distal villous hypoplasia, often occuring in placentas with long periods of fetal hypoxia, often co-occuring with other signs such as of accelerated villous maturation (39, 40). Signs of chorioamnionitis were found equally in our low and high-risk cohorts. However, in our high-risk cohort the presence of chorioamnionitis was associated with normal Z-scores, suggesting different pathological processes in these high-risk cases such as a more acute pathology associated with inflammation.

Automatic segmentation of the placenta has attracted reasonable interest mainly on anatomical data with the aim to perform volumetrics (13–15). Previous functional T2*-based studies have used manual segmentations of either individual slices (4, 6, 9) or the entire organ (8, 12). The presented study allows automatic segmentation of the entire placenta, but does not provide exact volumetrics due the distortions and inter-slice motion present in the Multi-Echo Gradient Echo data.

The aim of this pipeline was to establish a fully automatic assessment, close to the data and accessible throughout the processing pipeline. However, this could be expanded with further post-processing steps. These include placental 3D reconstruction, eg from orthogonal slice stacks or multiple dynamics as has been recently proposed for T2* data for the brain (41) and the placenta (28) or inter-slice motion correction techniques (42) to improve the 3D continuity of the data thus enabling 3D patch-based segmentation and/or placental flattening techniques (16, 43), resulting in representations in a common coordinate system.

This study deliberately proposes a pipeline built explicitly to be close to the original and wide-spread 2D multi-slice acquisition which makes it extendable to other applications of quantification of T2* data. It provides the full organ segmentation for all acquired slices and this can be expanded at any stage by reconstruction and registration approaches. Similarly, the proposed maturation quantification method is independent of the actual measurement method of the T2* values. For the evaluation performed here, mean T2* over the whole organ was chosen as this is the most widely used measure. It thus provides excellent validation of the segmentation and the new maturation assessment concept: The obtained mean T2* values as a function of age matches observations in the recent literature (4, 9). Further measures of interest for placental characterisation, including spatial information as previously proposed histogram based measures and texture analysis (8, 44, 45), can be included in future work. Another possible directly supported step would be the use of a convolutional neural network on the imaging data itself to map image properties against age instead of the mean T2* values.

The available data including comprehensive clinical information about maternal and fetal outcome has allowed differentiation between low- and high-risk cohorts, with evidence to support placental insufficiency in the latter. The low-risk cohort is crucial to train the prediction algorithm as it allows the reasonable assumption that the chronological age is an unbiased estimator of biological age. The availability of birth weight, gestation at birth and histopathology assessment results is essential to evaluate the acquired scores.

The segmentation U-net training requires a, for clinical settings, relatively large training set. The data used in this study was collected from a large scale study and presents over 100 datasets from in-vivo placental MRI scans together with comprehensive outcome information allowing the creation of a well characterized normal cohort, which is one of the largest such data collections. Rigorous selection of control cases was performed, only considering cases with complete outcome and no history of fetal or maternal complications (see section ‘Data’). However, for the high-risk cohort with suspected ‘placental insufficiency’, cases with either PE or fetal growth restriction have been analysed jointly to reach sufficient numbers. It is appreciated though that placental pathology may be different in these two groups and also between cases with early versus late onset PE. The PE cases were explored in a previous publication (26). The high-risk cohort in the 1.5T data was more uniform in the pathology as all were clinically diagnosed with PE, which was well reflected in the clearer separation in Z-scores. The available datasets are not equally distributed over GA with a specifically reduced occurrence before 22 weeks and most notably after 36 weeks. This both influences the ability of the segmentation U-net to identify placental tissue in later pregnancy (see decay in Dice coefficients over GA) and the relatively wide credibility interval of the Gaussian Process model for representations with lower mean T2* as would normally occur in later GA and in high-risk cases with later confirmed placental insufficiency. In addition, recruiting and scanning cases with suspected placental insufficiency is constrained by the challenges of clinical instability, need for urgent early delivery and the availability of scan slots. The reduced Dice scores for this cohort might again result from a lack of training samples for such placentas. The strength of the proposed pipeline is that it keeps the clinician in the loop and allows manual intervention via access to the T2* maps and segmentation masks.

Finally, if the achieved maturation marker is to be used in longitudinal studies to evaluate e.g the influence of treatments, its robustness to track individual pregnancies over time needs to be established with longitudinal data.

This study proposes a comprehensive, automatic pipeline to evaluate the maturation of a placenta in vivo from a short T2* MRI scan. Further factors which could be studied using this suggested pipeline include the effects and alteration of maturation in prolonged pregnancies or gestational diabetes. Inclusion into the clinical workflow is facilitated by the ease of use and the transparent steps employed.

## Data Availability

Medical imaging data is not made publicly available. Derived data and Gaussian Process analysis source code are made available.

https://osf.io/fbcsa/?view_only=324ffb5712f545aa8885f11261615857

## Acknowledgments

This work received funding from the NIH Human Placenta Project grant [1U01HD087202-01] European Research Council under the European Union’s Seventh Framework Programme [FP7/20072013]/ERC grant agreement no. [319456] (dHCP project), and was supported by the Wellcome EPSRC Centre for Medical Engineering at Kings College London [WT 203148/Z/16/Z], Wellcome Trust Sir Henry Wellcome Fellowship and Open Science Enrichment award, MRC strategic grant [MR/K006355/1] and by the National Institute for Health Research (NIHR) Biomedical Research Centre based at Guy’s and St Thomas’ NHS Foundation Trust and King’s College London. The views expressed are those of the authors and not necessarily those of the NHS, the NIHR or the Department of Health. The authors thank all involved radiographers, research midwifes and participating mothers for their invaluable help.

## Credit author statement

M Pietsch: Methodology, Writing, Software, Supervision; A Ho: Data curation, Investigation; A Bardanzellu: Investigation; A Zeidan: Investigation; L Chappell: Investigation, Supervision, Resources; J Hajnal: Supervision, Resources; Funding; M Rutherford: Supervision, Resources, Funding, Conceptualization; J Hutter: Conceptualization, Methodology, Writing, Software, Funding, Supervision. All authors read and approved the manuscript.

## Supplementary Material

**Supplementary Figure 8.**
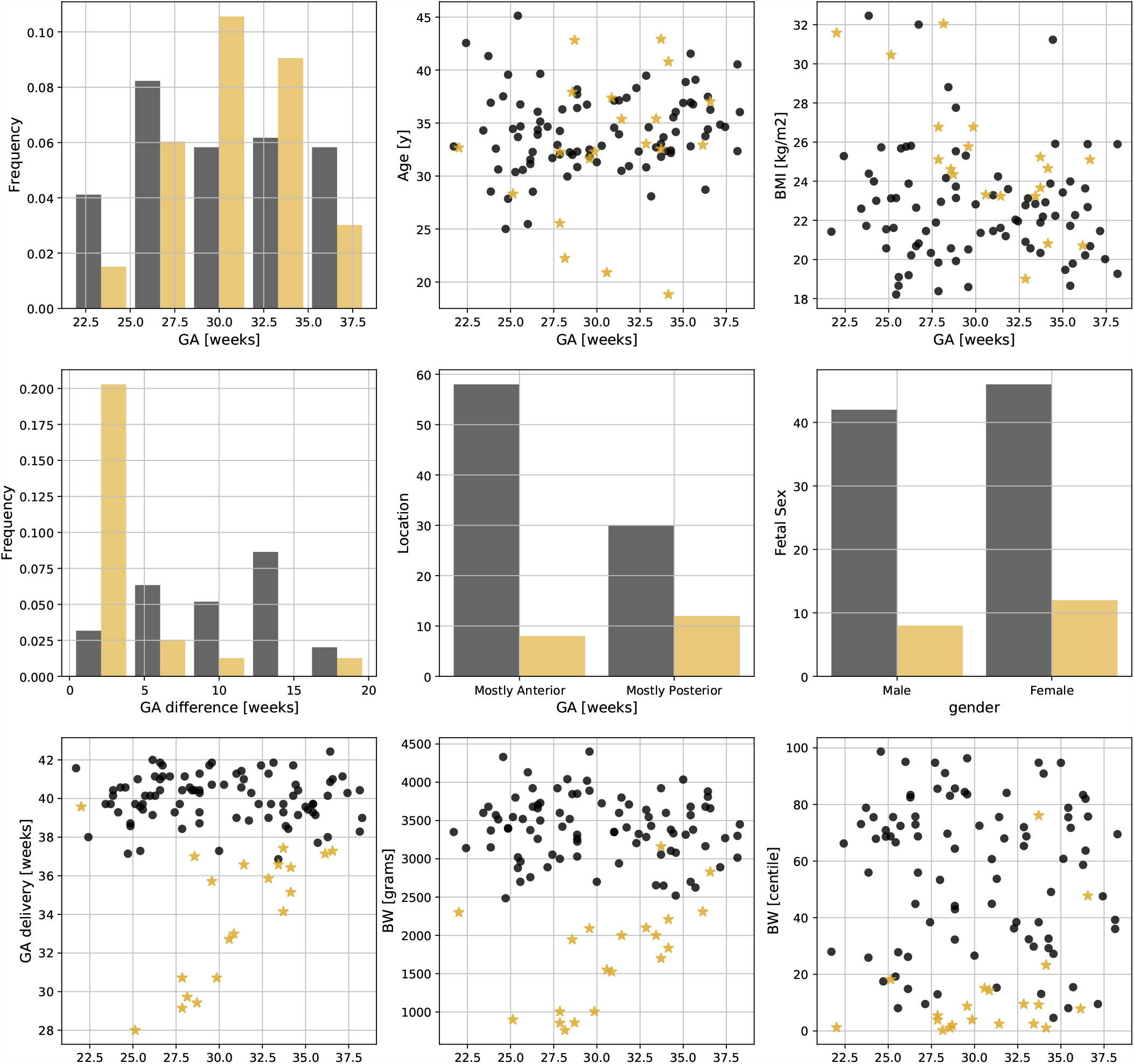
The demographics of the cohort are graphically illustrated with the control cohort in gray and the abnormal cohort in mustard. (a) Distribution of the GA at the time of scan, maternal age (b), and (c) BMI at the time of scan, distribution of the difference from GA at scan to GA at birth (d), placental location (e) and fetal sex (f) distribution. Finally, gestation at birth (g), birth weight (BW) in gram (h) and birth weight centile (i).

**Supplementary Figure 9.**
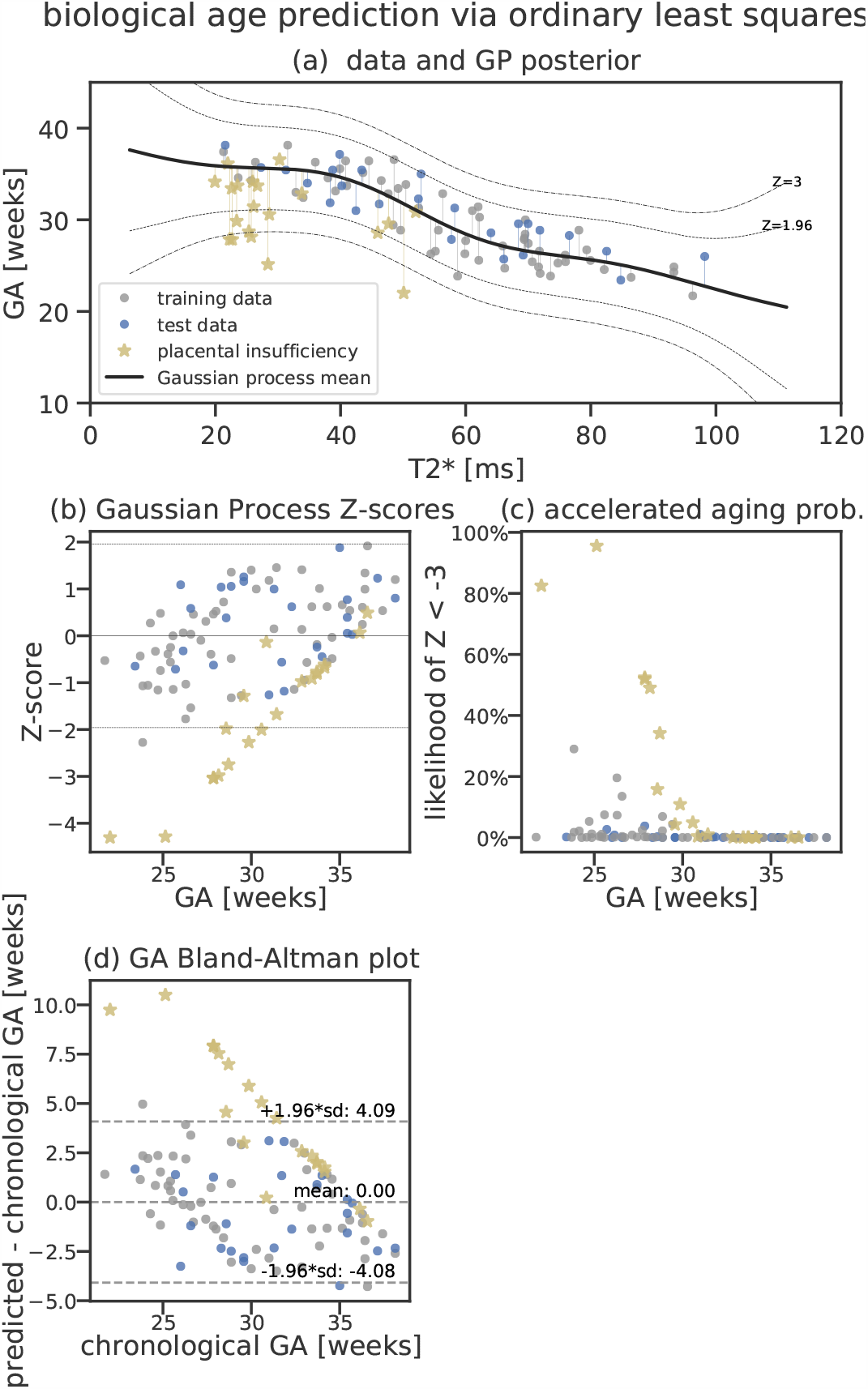
Ordinary Least Squares Gaussian Process results. This fit was performed with identical initial hyperparameters, fitting procedure and data as in Fig. 5 but using an ordinary least squares fit instead of the total least squares projection. Here, Z-scores are derived from the difference between chronological and predicted biological GA (b). The clear age-trend in the residuals for training and testing data indicate that an error-in-variable model is required.

**Supplementary Figure 10.**
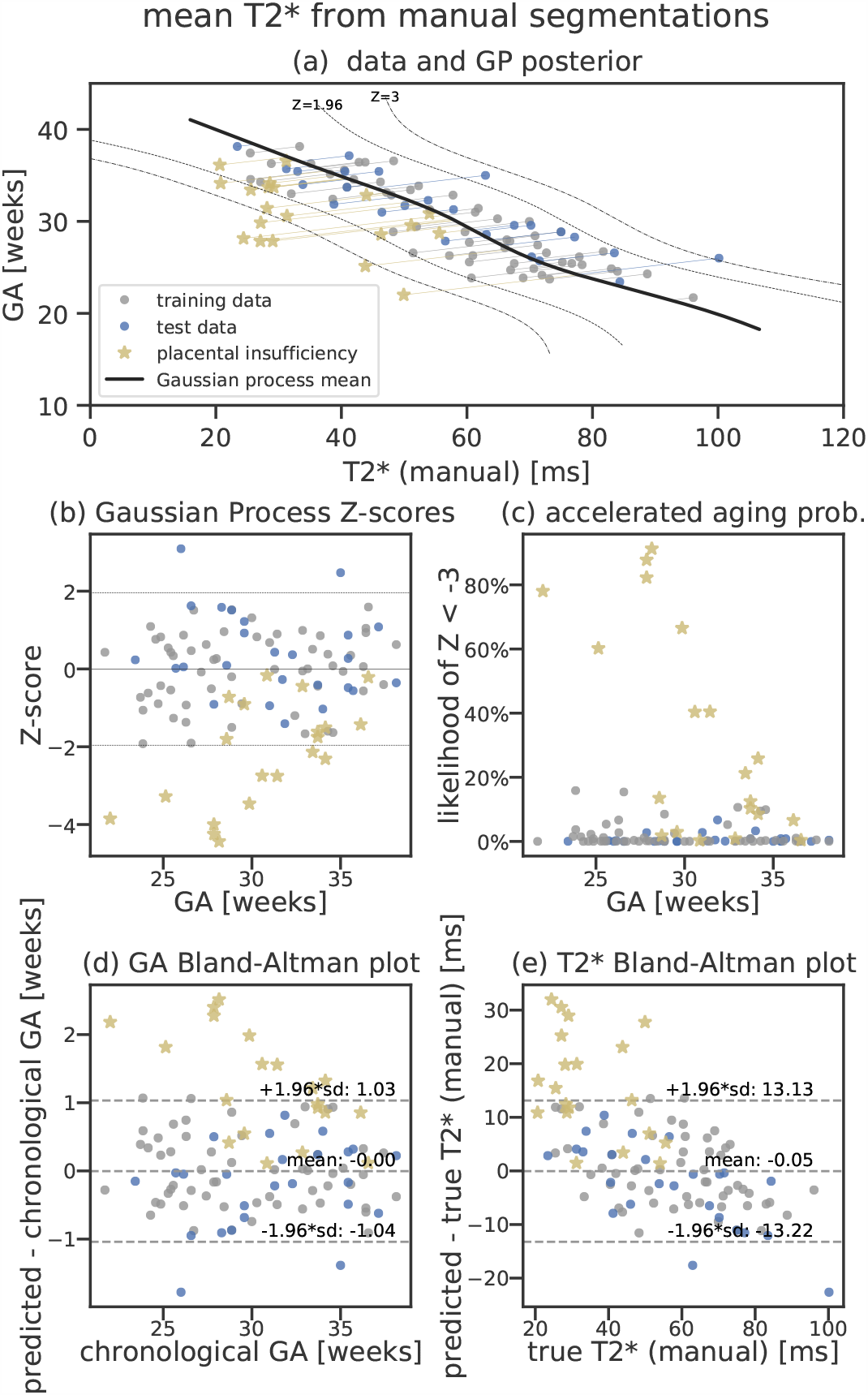
Equivalent of Fig. 5 using manual segmentations for T2* estimation.

**Supplementary Figure 11.**
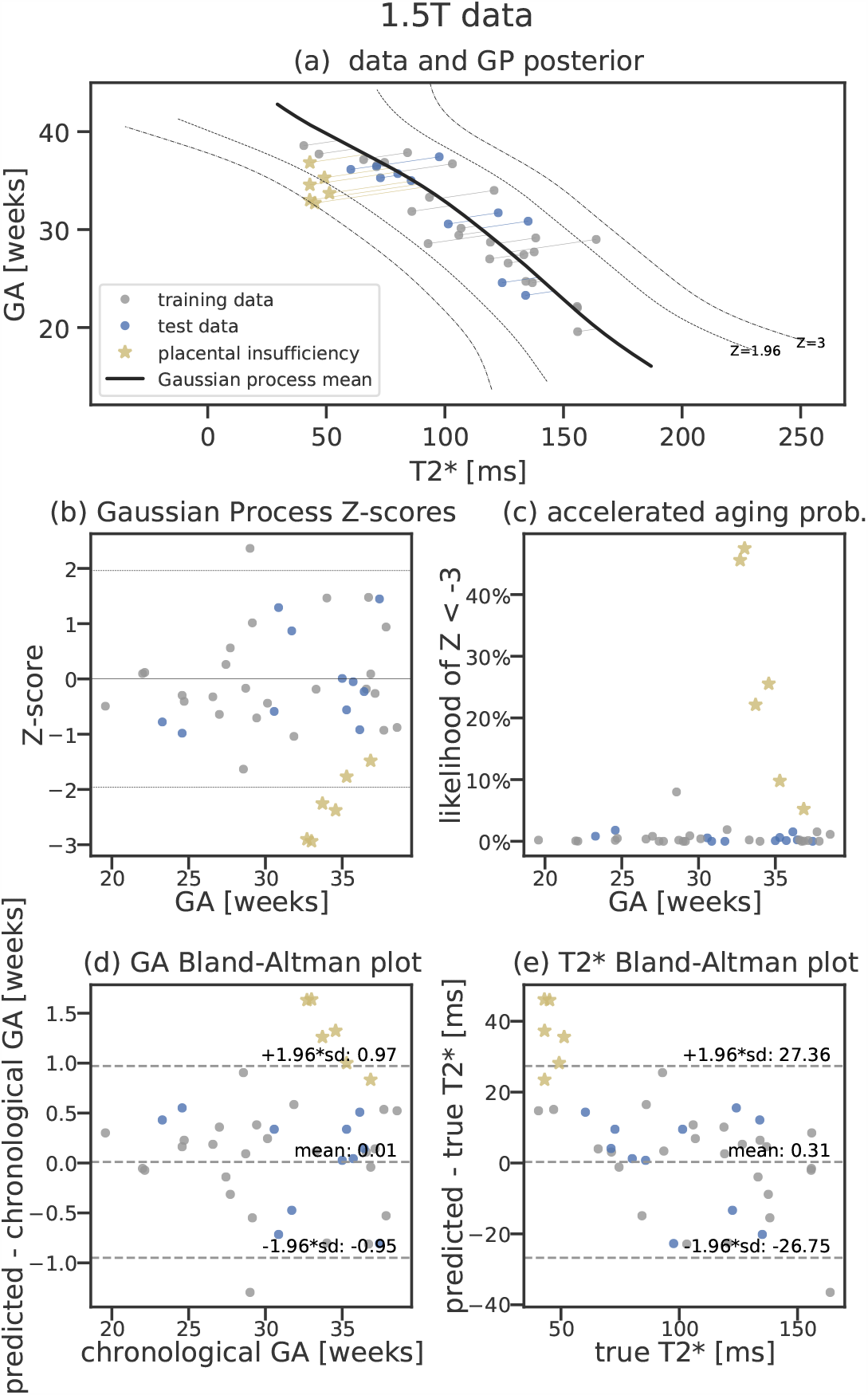
Equivalent of Fig. 5 using 1.5T data.

## Footnotes

1 https://github.com/MIC-DKFZ/batchgenerators

## References

1. Thomas Stallmach, Gundula Hebisch, Karin Meier, Joachim W. Dudenhausen, and Martin Vogel. Rescue by birth: Defective placental maturation and late fetal mortality. Obstetrics and Gynecology, 97(4):505–509, apr 2001. ISSN 0029-7844. doi: 10.1016/S0029-7844(00)01208-4.

2. Samantha J. Benton, Katherine Leavey, David Grynspan, Brian J. Cox, and Shannon A. Bainbridge. The clinical heterogeneity of preeclampsia is related to both placental gene expression and placental histopathology. American Journal of Obstetrics and Gynecology, 219(6):604.e1–604.e25, Dec 2018. ISSN 1097-6868. doi: 10.1016/j.ajog.2018.09.036.

3. Gitta Turowski and Martin Vogel. Re-view and view on maturation disorders in the placenta. APMIS, 126(7):602–612, jul 2018. ISSN 0903-4641. doi: 10.1111/apm.12858.

4. A Sørensen, D Peters, E Fründ, G Lingman, O Christiansen, and N Uldbjerg. Changes in human placental oxygenation during maternal hyperoxia estimated by blood oxygen level-dependent magnetic resonance imaging (BOLD MRI). Ultrasound in Obstetrics & Gynecology, 42(3):310–4, 2013. ISSN 1469-0705. doi: 10.1002/uog.12395.

5. Emma Ingram, David Morris, Josephine Naish, Jenny Myers, and Edward Johnstone. MR Imaging Measurements of Altered Placental Oxygenation in Pregnancies Complicated by Fetal Growth Restriction. Radiology, 285(3):953–960, dec 2017. doi: 10.1148/radiol.2017162385.

6. M. Sinding, D. A. Peters, J. B. Frøkjaer, O. B. Christiansen, A. Petersen, N. Uldbjerg, and A Sørensen. Placental magnetic resonance imaging T2*measurements in normal pregnancies and in those complicated by fetal growth restriction. Ultrasound in Obstetrics & Gynecology, 47(6):748–754, jun 2016. ISSN 0960-7692. doi: 10.1002/uog.14917.

7. Paddy J. Slator, Jana Hutter, Marco Palombo, Laurence H. Jackson, Alison Ho, Eleftheria Panagiotaki, Lucy C. Chappell, Mary A. Rutherford, Joseph V. Hajnal, and Daniel C. Alexander. Combined diffusion-relaxometry MRI to identify dysfunction in the human placenta. Magnetic Resonance in Medicine, 82(1):95–106, oct 2019. ISSN 1522-2594. doi: 10.1002/mrm.27733.

8. Jana Hutter, Paddy J. Slator, Laurence Jackson, Ana Dos Santos Gomes, Alison Ho, Lisa Story, Jonathan O’Muircheartaigh, Rui P. A. G. Teixeira, Lucy C. Chappell, Daniel C. Alexander, Mary A. Rutherford, and Joseph V. Hajnal. Multi-modal functional MRI to explore placental function over gestation. Magnetic Resonance in Medicine, sep 2018. doi: 10.1002/mrm.27447.

9. A. Sørensen, J. Hutter, M. Seed, P. E. Grant, and P. Gowland. T2* -weighted placental MRI: basic research tool or emerging clinical test for placental dysfunction? Ultrasound in Obstetrics and Gynecology, 55(3):293–302, mar 2020. ISSN 1469-0705. doi: 10.1002/uog.20855.

10. Esra Abaci Turk, Jie Luo, Natalie Copeland, Michelle Restrepo, Ata Turk Borjan Gagoski, Lawrence L. Wald, Elfar Adalsteinsson, Drucilla J. Roberts, Polina Golland, P. Ellen Grant, and William H. Barth Jr. Assessment of the effect of maternal posture on the placental oxygenation transport by means of BOLD MRI. In ISMRM 2018, 2018.

11. Neele S. Dellschaft, George Hutchinson, Simon Shah, Nia W. Jones, Chris Bradley, Lopa Leach, Craig Platt, Richard Bowtell, and Penny A. Gowland. The haemodynamics of the human placenta in utero. PLOS Biology, 18(5):1–21, 05 2020. doi: 10.1371/journal.pbio.3000676.

12. Esra Abaci Turk, S. Mazdak Abulnaga, Jie Luo, Jeffrey N. Stout, Henry A. Feldman, Ata Turk, Borjan Gagoski, Lawrence L. Wald, Elfar Adalsteinsson, Drucilla J. Roberts, Carolina Bibbo, Julian N. Robinson, Polina Golland, P. Ellen Grant, and William H. Barth. Placental MRI: Effect of maternal position and uterine contractions on placental BOLD MRI measurements. Placenta, 95:69–77, jun 2020. ISSN 1532-3102. doi: 10.1016/j.placenta.2020.04.008.

13. Guotai Wang, Maria A. Zuluaga, Rosalind Pratt, Michael Aertsen, Anna L. David, Jan Deprest, Tom Vercauteren, and Sebastien Ourselin. Slic-seg: Slice-by-slice segmentation propagation of the placenta in fetal MRI using one-plane scribbles and online learning. In Lecture Notes in Computer Science (including subseries Lecture Notes in Artificial Intelligence and Lecture Notes in Bioinformatics), volume 9351, pages 29–37. Springer Verlag, 2015. ISBN 9783319245737. doi: 10.1007/978-3-319-24574-4_4.

14. Amir Alansary, Konstantinos Kamnitsas, Alice Davidson, Rostislav Khlebnikov, Martin Rajchl, Christina Malamateniou, Mary Rutherford, Joseph V. Hajnal, Ben Glocker, Daniel Rueckert, and Bernhard Kainz. Fast fully automatic segmentation of the human placenta from motion corrupted MRI. In Lecture Notes in Computer Science (including subseries Lecture Notes in Artificial Intelligence and Lecture Notes in Bioinformatics), volume 9901 LNCS, pages 589–597. Springer Verlag, 2016. ISBN 9783319467221. doi: 10.1007/978-3-319-46723-8_68.

15. Mo Han, Yuwei Bao, Ziyan Sun, Shiping Wen, Liming Xia, Jingyang Zhao, Junfeng Du, and Zheng Yan. Automatic Segmentation of Human Placenta Images with U-Net. IEEE Access, 7:180083–180092, 2019. ISSN 2169-3536. doi: 10.1109/ACCESS.2019.2958133.

16. H. Miao, G. Mistelbauer, A. Karimov, A. Alansary, A. Davidson, D.F.A. Lloyd, M. Damodaram, L. Story, J. Hutter, J.V. Hajnal, M. Rutherford, B. Preim, B. Kainz, and M.E. Groller. Placenta maps: In utero placental health assessment of the human fetus. IEEE Transactions on Visualization and Computer Graphics, 23(6), 2017. ISSN 1077-2626. doi: 10.1109/TVCG.2017.2674938.

17. Gordon N. Stevenson, Sally L. Collins, Jane Ding, Lawrence Impey, and J. Alison Noble. 3-D Ultrasound Segmentation of the Placenta Using the Random Walker Algorithm: Reliability and Agreement. Ultrasound in Medicine and Biology, 41(12):3182–3193, ec 2015. ISSN 1879-291X. doi: 10.1016/j.ultrasmedbio.2015.07.021.

18. Marianne Sinding, David A. Peters, Jens B. Frøkjær, Ole B. Christiansen, Astrid Petersen, Niels Uldbjerg, and Anne Sørensen. Prediction of low birth weight: Comparison of placental T2*estimated by MRI and uterine artery pulsatility index. Placenta, 49:48–54, jan 2017. ISSN 0143-4004. doi: 10.1016/J.PLACENTA.2016.11.009.

19. Marianne Sinding, David A. Peters, Sofie S. Poulsen, Jens B. Frøkjær, Ole B. Christiansen, Astrid Petersen, Niels Uldbjerg, and Anne Sørensen. Placental baseline conditions modulate the hyperoxic BOLD-MRI response. Placenta, 61:17–23, jan 2018. doi: 10.1016/J.PLACENTA.2017.11.002.

20. Tess Armstrong, Dapeng Liu, Thomas Martin, Rinat Masamed, Carla Janzen, Cass Wong, Teresa Chanlaw, Sherin U. Devaskar, Kyunghyun Sung, and Holden H. Wu. 3D R2*Mapping of the Placenta During Early Gestation Using Free-Breathing Multiecho Stack-of-Radial MRI at 3T. Journal of Magnetic Resonance Imaging, aug 2018. doi: 10.1002/jmri.26203.

21. Carl Edward Rasmussen. Gaussian Processes in machine learning. Lecture Notes in Computer Science (including subseries Lecture Notes in Artificial Intelligence and Lecture Notes in Bioinformatics), 3176:63–71, 2004. ISSN 0302-9743. doi: 10.1007/978-3-540-28650-9_4.

22. James H. Cole and Katja Franke. Predicting Age Using Neuroimaging: Innovative Brain Ageing Biomarkers, dec 2017. ISSN 1878-108X.

23. J. H. Cole, S. J. Ritchie, M. E. Bastin, M.C. Valdés Hernández, S. Muñoz Maniega, N. Royle, J. Corley, A. Pattie, S. E. Harris, Q. Zhang, N. R. Wray, P. Redmond, R. E. Marioni, J. M. Starr, S. R. Cox, J. M. Wardlaw, D. J. Sharp, and I. J. Deary. Brain age predicts mortality. Molecular Psychiatry, 23(5):1385–1392, may 2018. ISSN 1476-5578. doi: 10.1038/mp.2017.62.

24. Jason Steffener, Christian Habeck, Deirdre O’Shea, Qolamreza Razlighi, Louis Bherer, and Yaakov Stern. Differences between chronological and brain age are related to education and self-reported physical activity. Neurobiology of Aging, 40:138–144, apr 2016. ISSN 1558-1497. doi: 10.1016/j.neurobiolaging.2016.01.014.

25. Aris T Papageorghiou, Eric O Ohuma, Douglas G Altman, Tullia Todros, Leila Cheikh Ismail, Ann Lambert, Yasmin A Jaffer, Enrico Bertino, Michael G Gravett, Manorama Purwar, et al. International standards for fetal growth based on serial ultrasound measurements: The Fetal Growth Longitudinal Study of the INTERGROWTH-21st Project. The Lancet, 384(9946):869–879, sep 2014. ISSN 1474-547X. doi: 10.1016/S0140-6736(14)61490-2.

26. Alison E.P. Ho, Jana Hutter, Laurence H. Jackson, Paul T. Seed, Laura McCabe, Mudher Al-Adnani, Andreas Marnerides, Simi George, Lisa Story, Joseph V. Hajnal, Mary A. Rutherford, and Lucy C. Chappell. T2s Placental Magnetic Resonance Imaging in Preterm Preeclampsia: An Observational Cohort Study. Hypertension, pages 1523–1531, 2020. ISSN 1524-4563. doi: 10.1161/HYPERTENSIONAHA.120.14701.

27. Francesc Figueras and Eduard Gratacós. Update on the diagnosis and classification of fetal growth restriction and proposal of a stage-based management protocol. Fetal diagnosis and therapy, 36(2):86–98, 2014. doi: 10.1159/000357592.

28. Uus Alena, Johannes K. Steinweg, Alison Ho, Laurence H. Jackson, Joseph V. Hajnal, Mary A. Rutherford, Maria Deprez, and Jana Hutter. Deformable Slice-to-Volume Registration for Reconstruction of Quantitative T2*Placental and Fetal MRI. In LNCS, ASMUS and PIPPI Proceedings, 2020.

29. Fabian Isensee Paul F. Jäger, Simon A. A. Kohl, Jens Petersen, and Klaus H. Maier-Hein. Automated Design of Deep Learning Methods for Biomedical Image Segmentation. preprint, apr 2019.

30. Fabian Isensee, Paul Jäger, Jakob Wasserthal, David Zimmerer, Jens Petersen, Simon Kohl, Justus Schock, Andre Klein, Tobias Roß, Sebastian Wirkert, Peter Neher, Stefan Dinkelacker, Gregor Köhler, and Klaus Maier-Hein. batchgenerators -a python framework for data augmentation. jan 2020. doi: 10.5281/ZENODO.3632567.

31. Diederik P. Kingma and Jimmy Lei Ba. Adam: A method for stochastic optimization. In 3rd International Conference on Learning Representations, ICLR 2015 - Conference Track Proceedings. International Conference on Learning Representations, ICLR, dec 2015.

32. Lee R. Dice. Measures of the Amount of Ecologic Association Between Species. Ecology, 26(3):297–302, jul 1945. ISSN 0012-9658. doi: 10.2307/1932409.

33. F Pedregosa, G Varoquaux, A Gramfort, V Michel,B Thirion, O Grisel, M Blondel, P Prettenhofer, R Weiss, V Dubourg, J Vanderplas, A Passos, D Cournapeau, M Brucher, M Perrot, and E Duchesnay. Scikit-learn: Machine Learning in Python. Journal of Machine Learning Research, 12:2825–2830, 2011.

34. Chi-Lun Cheng and John W Van Ness. Statistical regression with measurement error. John Wiley & Sons, London New York, 1999. ISBN 0340614617.

35. Andrew Chesher. The effect of measurement error. Biometrika, 78 (3):451–462, 1991. doi: 10.1093/biomet/78.3.451.

36. Sabine Van Huffel. Total least squares and errors-in-variables modeling: Bridging the gap between statistics, computational mathematics and engineering. In COMPSTAT 2004—Proceedings in Computational Statistics, pages 539–555. Springer, 2004.

37. Susanne M Schennach. Recent advances in the measurement error literature. Annual Review of Economics, 8:341–377, 2016. doi: 10.1146/annurev-economics-080315-015058.

38. Christina L Hennig, Jessie Childs, Aamer Aziz, and Ann Quinton. The effect of increased maternal body habitus on image quality and ability to identify fetal anomalies at a routine 18-20-week morphology ultrasound scan: a narrative review. Sonography, 6(4):191–202, 2019. doi: 10.1002/sono.12202.

39. W Tony Parks and Janet M Catov. The Placenta as a Window to Mater nal Vascular Health. Obstetrics and Gynecology Clinics of NA, 47:17–28, 2020. doi: 10.1016/j.ogc.2019.10.001.

40. Linda M. Ernst. Maternal vascular malperfusion of the placental bed. APMIS, 126(7):551–560, jul 2018. ISSN 0903-4641. doi: 10.1111/apm.12833.

41. Anna I. Blazejewska, Sharmishtaa Seshamani, Susan K. McKown, Jason S. Caucutt, Manjiri Dighe, Christopher Gatenby, and Colin Studholme. 3D in utero quantification of T2*relaxation times in human fetal brain tissues for age optimized structural and functional MRI. Magnetic Resonance in Medicine, 78(3):909–916, sep 2017. ISSN 0740-3194. doi: 10.1002/mrm.26471.

42. Jie Luo, Esra Abaci Turk, Carolina Bibbo, Borjan Gagoski, Drucilla J. Roberts, Mark Vangel, Clare M. Tempany-Afdhal, Carol Barnewolt, Judy Estroff, Arvind Palanisamy, William H. Barth, Chloe Zera, Norberto Malpica, Polina Golland, Elfar Adalsteinsson, Julian N. Robinson, and Patricia Ellen Grant. In Vivo Quantification of Placental Insufficiency by BOLD MRI: A Human Study. Scientific Reports, 7 (1):3713, dec 2017. doi: 10.1038/s41598-017-03450-0.

43. S. Mazdak Abulnaga, Esra Abaci Turk, Mikhail Bessmeltsev, P. Ellen Grant, Justin Solomon, and Polina Golland. Placental flattening via volumetric parameterization. In Lecture Notes in Computer Science (including subseries Lecture Notes in Artificial Intelligence and Lecture Notes in Bioinformatics), volume 11767 LNCS, pages 39–47. Springer, oct 2019. ISBN 9783030322502. doi: 10.1109/TMI.2020.2974844.

44. Jamie O. Lo, Victoria H. J. Roberts, Matthias C. Schabel, Xiaojie Wang, Terry K. Morgan, Zheng Liu, Colin Studholme, Christopher D. Kroenke, and Antonio E. Frias. Novel Detection of Placental Insufficiency by Magnetic Resonance Imaging in the Nonhuman Primate. Reproductive Sciences, 25(1):64–73, jan 2018. doi: 10.1177/1933719117699704.

45. Alec J. Hirsch, Victoria H. J. Roberts, Peta L. Grigsby, Nicole Haese, Matthias C. Schabel, Xiaojie Wang, Jamie O. Lo, Zheng Liu, Christopher D. Kroenke, Jessica L. Smith, Meredith Kelleher, Rebecca Broeckel, Craig N. Kreklywich, Christopher J. Parkins, Michael Denton, Patricia Smith, Victor DeFilippis, William Messer, Jay A. Nelson, Jon D. Hennebold, Marjorie Grafe, Lois Colgin, Anne Lewis, Rebecca Ducore, Tonya Swanson, Alfred W. Legasse, Michael K. Axthelm, Rhonda MacAllister, Ashlee V. Moses, Terry K. Morgan, Antonio E. Frias, and Daniel N. Streblow. Zika virus infection in pregnant rhesus macaques causes placental dysfunction and immunopathology. Nature Communications, 9(1):263, dec 2018. doi: 10.1038/s41467-017-02499-9.

